# Investigating Gender Disparities in Ophthalmology Departments at Medical Schools in Japan

**DOI:** 10.1101/2024.09.01.24312900

**Authors:** Akemi Iwasaki, Naoko Kato, Yuka Morita, Hiromi Onouchi, Mariko Itakura, Keiko Kunimi, Yoichi Manabe

**Author notes:** These authors contributed equally to this work (Akemi Iwasaki, Naoko Kato).

## Abstract

This comprehensive study investigates the gender distribution of ophthalmologists in academic positions in Japanese medical schools. Data were collected from the websites of all ophthalmology departments and affiliated hospitals from November 1-15, 2023. Faculty gender was identified using first names and photographs, and where unclear, further verification was sought from the Ministry of Health, Labour, and Welfare or known contacts. The survey included 1,574 faculty members: 453 females (28.8%) and 1,121 males (71.2%). The representation of females varied significantly across different academic ranks: 9.9% were professors, 21.9% associate professors, 39.5% assistant professors, and 36.7% research associates. A significant gender disparity was observed, with males being 4.41 times more likely to become professors than females (p < 0.001). Conversely, females were more likely to hold research associate positions than males (Odds Ratio: 0.51, p < 0.001). This study highlights a male predominance in senior academic positions within ophthalmology departments in Japanese medical schools.

## Introduction

The number of female physicians has been increasing worldwide in recent years. The global percentage of female physicians has now reached 50%, with particularly higher representation in Eastern Europe, South America, and Africa [1]. Despite the increasing prevalence of female physicians, there is evidence to suggest that women continue to be underrepresented in leadership roles, even in western countries [2-6]. According to recent data, male physicians are approximately 2.77 times more likely to hold a full-time professorship compared to their female counterparts [7].

In Japan, female physicians account for only slightly more than 20.5%, which is the lowest among the OECD countries [8]. Out of those, the percentage of females in the field of ophthalmology is relatively high [9]. However, the specific numbers of male and female ophthalmologists working with academic titles at the ophthalmology departments in Japanese medical universities have not been reported to the extent of our knowledge. Therefore, we investigated the gender disparity of ophthalmologists with the academic positions in Japanese medical schools in this study.

## Methods

We conducted a comprehensive survey assessing the gender distribution of faculty members in the departments of ophthalmology at medical schools in Japan. This study was approved by the Otaki Eye Clinic Ethics Committee (Committee Number: 21000020, Application Number: 16), and the need for consent was waived by the ethics committee. Data were collected from the websites of all ophthalmology departments and their affiliated hospitals during November 1-15, 2023. Faculty gender was primarily determined from first names and photographs, where available. Where gender was not identifiable from the website, we consulted the search engine of the Ministry of Health, Labour, and Welfare (MHLW) or reached out to known contacts for clarification.

Faculty members were categorized into four academic ranks: Professors, Associate Professors, Assistant Professors, and Research Associates. Chief professor, professor, clinical professor, treatment professor, hospital professor, non-tenured professor, project professor, endowed chair, professor emeritus, visiting professor, and visiting clinical professor were classified as Professors. Associate professor, clinical associate professor, treatment associate professor, hospital associate prof, non-tenured associate professor, associate professor in hospital, endowed associate chair, associate professor emeritus, visiting associate professor, and visiting clinical associate professor were categorized as Associate Professors. Senior lecturer, non-tenured lecturer, lecturer in the school of medicine, clinical lecturer, hospital lecturer, in-hospital lecturer, lecturer in the school, lecturer in the department, concurrent lecturer, lecturer in the graduate school, associate lecturer, specified lecturer, and part-time lecturer were classified as Assistant Professors. Assistant professor, special assistant professor, clinical assistant professor, fixed-term assistant professor, intramural assistant professor, hospital assistant professor, specified assistant professor, in-hospital assistant professor, clinical assistant professor, research assistant, medical school assistant, and medical assistant were categorized as Research Associates.

Statistical analyses were performed using JMP12 software (SAS Institute Inc. USA). We considered p-values < 0.05 as statistically significant. Our analytical approach included the use of odds ratios (OR) and chi-squared tests to examine gender disparities across the academic roles.

## Results

The study encompassed 1,574 faculty members in 81 ophthalmology departments, including 453 (28.8%) females and 1,121 (71.2%) males.

### Gender Distribution by Academic Position

There were 272 professors, 146 associate professors, 488 assistant professors, and 668 research associates. The percentage of female representation among professors was 9.9%, while among associate professors it was 21.9%, among assistant professors it was 39.5%, and among research associates it was 36.7% (Table 1, Fig 1). The distribution of each academic rank was significantly different between genders (p < 0.001; Fig 1).

**Table 1.** Numbers and incidence of gender at each position.

|  | **Numbers**<br>**(Females; males)** | **Female ratio** | **Odds ratio** | **p-value** |
| --- | --- | --- | --- | --- |
| **Professors** | 27; 245 | 9.9% | 4.41 | <0.001 |
| **Associate professors** | 32; 114 | 21.9% | 1.49 | 0.058 |
| **Assistant professors** | 149; 339 | 30.5% | 0.88 | 0.332 |
| **Research associates** | 245; 423 | 36.7% | 0.51 | <0.001 |

**Fig 1.**
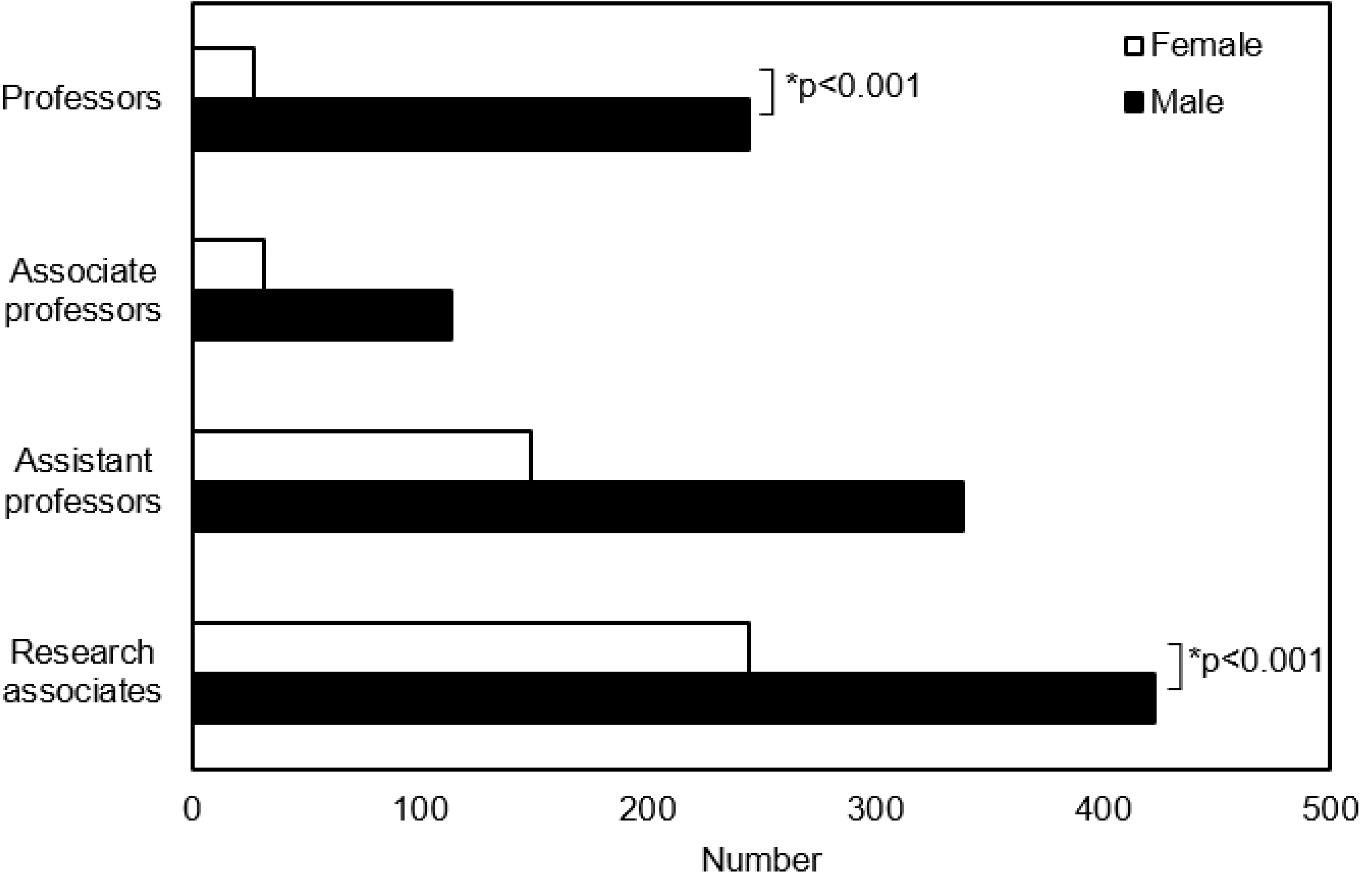
Gender distribution by academic positions. The number of each academic rank was significantly different between genders (p < 0.001). The odds of becoming professors were approximately 4.41 times higher in males than in females (Odds Ratio: 4.41, p < 0.001). Among all positions, females were more likely to occupy the role of research associates (Odds Ratio: 0.51, p < 0.001).

The odds of becoming professors were approximately 4.41 times higher for males than for females (p < 0.001), whereas females were more likely to occupy research associate positions than males (Odds Ratio: 0.51, p < 0.001; Table 1).

### Full-time and Part-time Jobs

Of the 1,574 members, 1,113 (70.7%) had full-time and 461 (29.3%) part-time jobs. Three hundred twenty-one (28.8%) were female and 792 (71.2%) were male among the full-time workers, and 132 (28.6%) were female and 329 (71.4%) were male among the part-time workers. There were no significant differences in the female ratio between full-time or part-time jobs (p = 0.983). The female ratio was also not significantly different between the full-time and part-time jobs in each academic position, except for the research associates (35.4% in full-time jobs and 53.2% in part-time jobs, p = 0.023; Fig 2).

**Fig 2.**
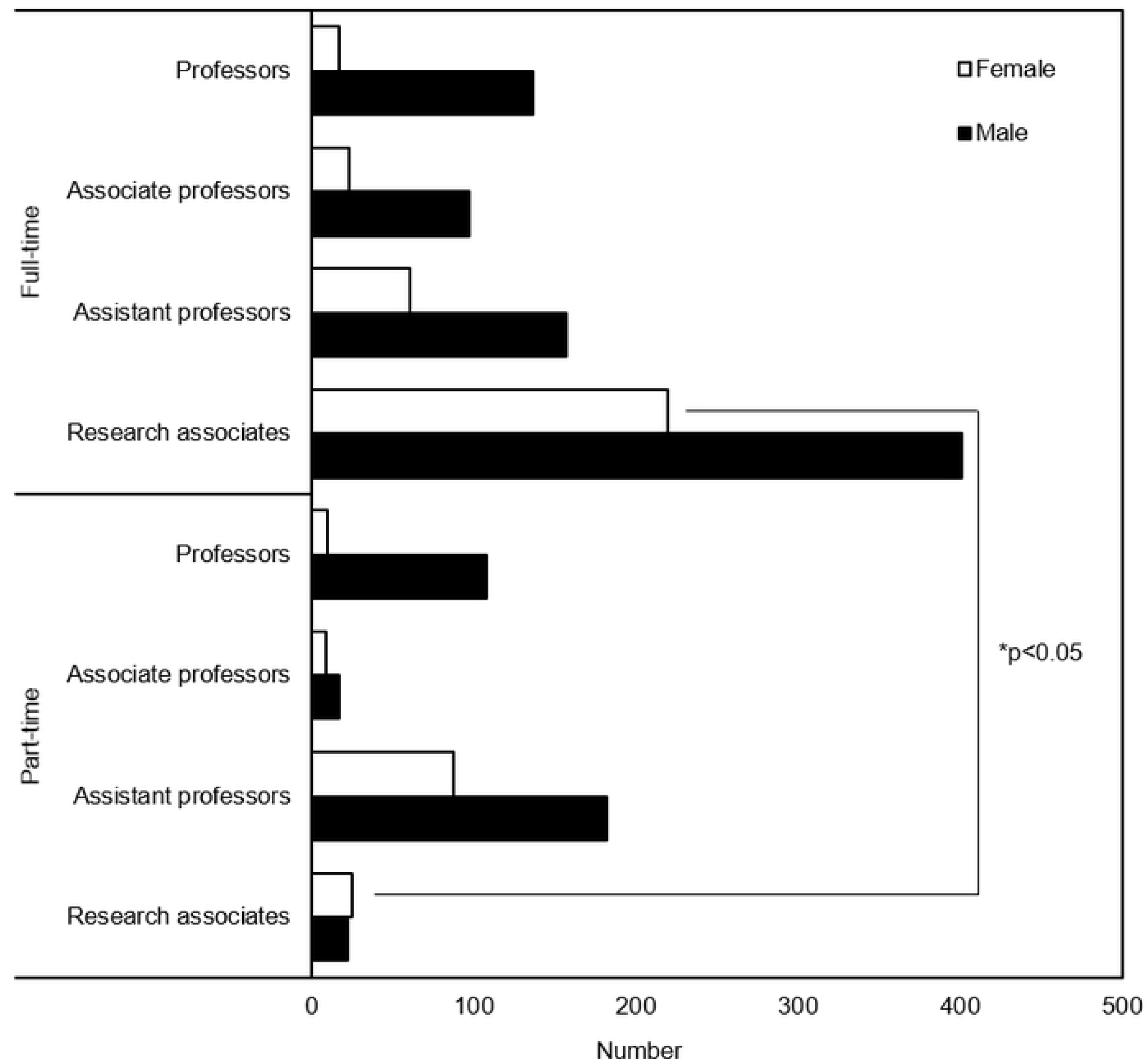
Differences of gender distribution by academic positions between full-time and part-time jobs. The real number of faculty members shows that women were mainly research associates, and they were less likely than men to hold higher academic posts in full-time positions. Part-time female assistant professors are notably abundant. The proportion of women among research associates was significantly higher in part-time positions (p = 0.02), whereas in other positions, there was no difference in gender ratio between full-time and part-time.

### Gender Differences in Faculty Composition Based on Chair Professor’s Gender

When we classified the faculty members according to the gender of the faculty’s chair professor, we observed a higher proportion of females in the institutes where the chair professors were female, compared to those with male chair professors (35.2% and 28.0%, respectively; p = 0.046; Table 2).

**Table 2.** Female ratio at each position based on chair professor’s gender.

|  | **Professors** | **Associate**<br>**professors** | **Assistant**<br>**professors** | **Research**<br>**associates** | **Total** |
| --- | --- | --- | --- | --- | --- |
| **Female chief professors** | 23.1% | 45.5% | 29.4% | 47.5% | 35.2% |
| **Male chief professors** | 7.7% | 20.0% | 30.7% | 35.6% | 28.0% |
| **p-value** | 0.0074 | 0.1133 | 0.9407 | 0.0877 | 0.046 |

## Discussion

The present study indicated a higher representation of males in academic professional roles, especially in senior positions within the ophthalmic department at university hospitals in Japan. In fact, males significantly outnumbered females, being 4.41 times more likely to become professors. In contrast, females were more likely to occupy research associate positions compared to their male counterparts.

According to a survey by the MHLW, the current number of female physicians in Japan is 67,671 (excluding clinical trainees without a specialty). Ophthalmology includes 5,364 (7.9%) female physicians and ranks fourth place among all specialties, following internal medicine, pediatrics, and dermatology [10]. Despite females accounting for 38.8% of ophthalmologists, the percentage of female professors in ophthalmology is only 9.9%, consistent with the percentage of female professors across all medical specialties in Japan [11]. These facts strongly suggest that achieving professorship is particularly challenging among the female ophthalmologists, who represent a significant presence in the field.

In Japan, it is customary for young medical school graduates to commence their careers as research associates. They then advance through the ranks of assistant professor, associate professor, and ultimately professor, based on their accomplishments in the fields of clinical practice and research. According to the current data, there is a decline in the ratio of females as academic positions become more elevated. The disparity in female representation cannot be attributed solely to the underrepresentation of females among older ophthalmologists. This is evident from the fact that the overall female ratio among ophthalmologists spans across generations, ranging from 31% (70-74 years old) to 47% (25-29 years old) [10].

A similar tendency was found in the gender ratio between part-time and full-time positions. In Japan, the designations of part-time professor, associate professor, and assistant professor are honorary titles granted to individuals who typically work in other medical institutions or clinics. Their primary role is to provide specialized outpatient services and/or deliver lectures to students. The probability of females becoming professors in both types of employment was reduced.

Conversely, the prevalence of females was only higher among part-time research associates compared to full-time employees. Part-time research associates in Japan often don’t have academic responsibilities, but the position offers employment opportunities for women unable to work full-time due to lifestyle changes [12-17]. Our previous research [18] indicated that many female ophthalmologists changed their places of employment and work styles after childbirth. This suggests that part-time research associate positions are being utilized to accommodating these complex circumstances.

A 2019 survey [19] by the MHLW on physicians’ working conditions reports that university hospitals have a higher percentage of physicians working over 60 hours per week compared to general hospitals. Specifically, the survey indicates that 44.2% of female physicians and 47.3% of male physicians in their twenties, 26.5% and 52.2% in their thirties, 21.5% and 43.9% in their forties, 24.2% and 36.4% in their fifties, and 15.1% and 20.4% in their sixties work these extended hours. Across all age groups, males exhibited a higher propensity for working extended hours, and the disparity between genders progressively intensified from the thirties onward. In Japanese society, working longer hours has historically been more highly rewarded [20-22], and physicians are no exception [23-24]. The present data strongly suggests that men may work longer hours instead of women, positioning themselves more favorably for career advancement.

The proportion of female medical students is currently increasing in Japan after an entrance examination gender discrimination scandal at Japanese medical schools [25-26], and the incidence of female physicians is expected to continue to rise. This situation calls for a shift in mindset. Physicians should also be evaluated based on their ability to achieve results, rather than solely focusing on the duration of their working hours.

The Japanese medical system will cease to function unless we reconsider the system of employment as soon as possible. Interestingly, institutes under the leadership of female chair professors exhibited a greater representation of female members in academic positions, in contrast to those under male chair professors. We consider that this could be one solution to the gender disparity in the ophthalmology departments in Japan. A higher representation of women in leadership roles could remove barriers for female ophthalmologists in academia, potentially resulting in a new evaluation and promotion system for all physicians in Japan.

An inherent limitation of this study is the exclusion of age, Ph.D. titles, marital status, and presence of children for the ophthalmologists as factors for analysis. Data collection was solely conducted through university hospital websites. The data of those without titles, graduate students, or those in female support slots could also not be collected. Further study is needed to clarify the gender disparities of promotion among Japanese ophthalmologists.

## Conclusions

This study revealed that positions within ophthalmology departments are predominantly occupied by males, particularly in higher positions. To overcome these issues, we need to clarify what aspects of the current culture and structure of Japanese ophthalmology departments are contributing to gender disparities.

## Data Availability

All relevant data are within the manuscript and its Supporting Information files.

## Acknowledgments

We express our gratitude to Mr. Ron Beaubien for his English language guidance.

## References

1. INDICATORS OECD. Health at a Glance 2021.

2. Blumenthal DM, Bergmark RW, Raol N, Bohnen JD, Eloy JA, Gray ST. Sex Differences in Faculty Rank Among Academic Surgeons in the United States in 2014. Ann Surg. 2018;268(2):193–200.

3. Jena AB, Khullar D, Ho O, Olenski AR, Blumenthal DM. Sex Differences in Academic Rank in US Medical Schools in 2014. Jama. 2015;314(11):1149–58.

4. Rochon PA, Davidoff F, Levinson W. Women in Academic Medicine Leadership: Has Anything Changed in 25 Years? Acad Med. 2016;91(8):1053–6.

5. Thibault GE. Women in Academic Medicine. Acad Med. 2016;91(8):1045–6.

6. Carr PL, Raj A, Kaplan SE, Terrin N, Breeze JL, Freund KM. Gender Differences in Academic Medicine: Retention, Rank, and Leadership Comparisons From the National Faculty Survey. Acad Med. 2018;93(11):1694–9.

7. Li B, Jacob-Brassard J, Dossa F, Salata K, Kishibe T, Greco E, et al. Gender differences in faculty rank among academic physicians: a systematic review and meta-analysis. BMJ Open. 2021;11(11):e050322.

8. Akazawa S, Fujimoto Y, Sawada M, Kanda T, Nakahashi T. Women Physicians in Academic Medicine of Japan. JMA J. 2022 Jul 15;5(3):289–297.

9. Yonemoto K, The female physician’s present condition in Japan : comparison with foreign countries. Doshisha policy and managemant review. 2012; 13 (2), 109–125.

10. Ministry of Health, Labour and Welfare. Overview of the 2022 Statistics on Physicians, Dentists, and Pharmacists. https://www.mhlw.go.jp/toukei/saikin/hw/ishi/22/index.html (accessed 2024-07-12).

11. Kono K, Watari T, Tokuda Y. Assessment of Academic Achievement of Female Physicians in Japan. JAMA Netw Open. 2020;3(7):e209957.

12. Suzuki S. Exhausting Physicians Employed in Hospitals in Japan Assessed by a Health Questionnaire. Sangyo Eiseigaku Zasshi. 2017;59(4):107–18.

13. Nomura K, Sato M, Tsurugano S, Yano E. Factors associated with working among female physicians in Japan. Nihon Koshu Eisei Zasshi. 2011;58(6):433–45.

14. Kaneto C, Toyokawa S, Inoue K, Kobayashi Y. Gender difference in physician workforce participation in Japan. Health Policy. 2009;89(1):115–23.

15. Nomura K, Yamazaki Y, Gruppen LD, Horie S, Takeuchi M, Illing J. The difficulty of professional continuation among female doctors in Japan: a qualitative study of alumnae of 13 medical schools in Japan. BMJ Open. 2015;5(3):e005845.

16. Izumi M, Nomura K, Higaki Y, Akaishi Y, Seki M, Kobayashi S, et al. Gender role stereotype and poor working condition pose obstacles for female doctors to stay in full-time employment: alumnae survey from two private medical schools in Japan. Tohoku J Exp Med. 2013;229(3):233–7.

17. Hashizume Y. Gender issues and Japanese family-centered caregiving for frail elderly parents or parents-in-law in modern Japan: from the sociocultural and historical perspectives. Public Health Nurs. 2000;17(1):25–31.

18. Kato N, Kojima T, Ouchi M, Nakamura T, Tokuda Y, Yakushiji T, et al. Gender-based differences in the job titles and lifestyles in the cataract and refractive surgery society in Japan. Medicine (Baltimore). 2023;102(40):e35216.

19. The 2019 Survey on the Working Conditions of Physicians. Available from: https://www.mhlw.go.jp/stf/newpage_05488.html.

20. Gicheva D. Working long hours and early career outcomes in the high-end labor market. Journal of Labor Economics. 2013;31(4):785–824.

21. Nemoto K. Long Working Hours and the Corporate Gender Divide in Japan. Gender, Work and Organization. 2013;20:512–27.

22. Kato T, Ogawa H, Owan H. Working Hours, Promotion and the Gender Gap in the Workplace. IZA DP No. 10454. 2016.

23. Hiyama T, Yoshihara M. New occupational threats to Japanese physicians: karoshi (death due to overwork) and karojisatsu (suicide due to overwork). Occup Environ Med. 2008;65(6):428–9.

24. Koike S, Wada H, Ohde S, Ide H, Taneda K, Tanigawa T. Working hours of full-time hospital physicians in Japan: a cross-sectional nationwide survey. BMC Public Health. 2024;24(1):164.

25. Watari T, Mizuno K, Sakaguchi K, Shimada Y, Tanimoto Y, Nakano Y, et al. Gender Inequality Improvement in Medical School Admissions in Japan. J Womens Health (Larchmt). 2024;33(3):339–44.

26. Dyer O. Tokyo medical school investigates claims that it rigged exam results to turn women away. Bmj. 2018;362:k3416

